# Multicenter Validation of a Machine Learning Algorithm for Diagnosing Pediatric Patients with Multisystem Inflammatory Syndrome and Kawasaki Disease

**DOI:** 10.1101/2022.02.07.21268280

**Authors:** Jonathan Y. Lam, Samantha C. Roberts, Chisato Shimizu, Emelio Bainto, Nipha Sivilay, Adriana H. Tremoulet, Michael A. Gardiner, John T. Kanegaye, Alexander H. Hogan, Juan C. Salazar, Sindhu Mohandas, Jacqueline R. Szmuszkovicz, Simran Mahanta, Audrey Dionne, Jane W. Newburger, Emily Ansusinha, Roberta L. DeBiasi, Shiying Hao, Xuefeng B. Ling, Harvey J. Cohen, Shamim Nemati, Jane C. Burns, the Pediatric Emergency Medicine Kawasaki Disease Research Group, the CHARMS Study Group

## Abstract

**Background:** Multisystem inflammatory syndrome in children (MIS-C) is a novel disease identified during the COVID-19 pandemic characterized by systemic inflammation following SARS-CoV-2 infection. Delays in diagnosing MIS-C may lead to more severe disease with cardiac dysfunction or death. Most pediatric patients recover fully with anti-inflammatory treatments, but early detection of MIS-C remains a challenge given its clinical similarities to Kawasaki disease (KD) and other acute childhood illnesses.

**Methods:** We developed KIDMATCH (<u>K</u>awasak<u>I</u> <u>D</u>isease vs <u>M</u>ultisystem Infl<u>A</u>mma<u>T</u>ory syndrome in <u>CH</u>ildren), a deep learning algorithm for screening patients for MIS-C, KD, or other febrile illness, using age, the five classical clinical KD signs, and 17 laboratory measurements prospectively collected within 24 hours of admission to the emergency department from 1448 patients diagnosed with KD or other febrile illness between January 1, 2009 and December 31, 2019 at Rady Children’s Hospital. For MIS-C patients, the same data was collected from 131 patients between May 14, 2020 to June 18, 2021 at Rady Children’s Hospital, Connecticut Children’s Hospital, and Children’s Hospital Los Angeles. We trained a two-stage model consisting of feedforward neural networks to distinguish between MIS-C and non MIS-C patients and then KD and other febrile illness. After internally validating the algorithm using 10-fold cross validation, we incorporated a conformal prediction framework to tag patients with erroneous data or distribution shifts, enhancing the model generalizability and confidence by flagging unfamiliar cases as indeterminate instead of making spurious predictions. We externally validated KIDMATCH on 175 MIS-C patients from 16 hospitals across the United States.

**Findings:** KIDMATCH achieved a high median area under the curve in the 10-fold cross validation of 0.988 [IQR: 0.98-0.993] in the first stage and 0.96 [IQR: 0.956-0.972] in the second stage using thresholds set at 95% sensitivity to detect positive MIS-C and KD cases respectively during training. External validation of KIDMATCH on MIS-C patients correctly classified 76/83 (2 rejected) patients from the CHARMS consortium, 47/50 (1 rejected) patients from Boston Children’s Hospital, and 36/42 (2 rejected) patients from Children’s National Hospital.

**Interpretation:** KIDMATCH has the potential to aid frontline clinicians with distinguishing between MIS-C, KD, and similar febrile illnesses in a timely manner to allow prompt treatment and prevent severe complications.

**Funding:** Eunice Kennedy Shriver National Institute of Child Health and Human Development, National Heart, Lung, and Blood Institute, Patient-Centered Outcomes Research Institute, National Library of Medicine

## Introduction

As the coronavirus disease (COVID-19) pandemic spread, reports of children with SARS-CoV-2-associated multisystem inflammatory conditions emerged^1–6^. Clinical features of this new disorder, named Multisystem Inflammatory Syndrome in Children (MIS-C), include fever, gastrointestinal symptoms, and elevated inflammatory markers^7^. Complications may include shock and multiorgan failure. According to the Centers for Disease Control and Prevention (CDC), a total of 5,973 MIS-C cases and 52 MIS-C deaths have been reported nationwide as of November 30, 2021^8^. Despite its low prevalence, MIS-C is a serious condition with potential to cause life-threatening illness, and the lack of a specific diagnostic test makes recognizing MIS-C a challenge.

Treatments for MIS-C include intravenous immunoglobulin (IVIG), corticosteroids, and anti-inflammatory biologic agents that rely on timely diagnosis of MIS-C to be most effective^9^. Kawasaki disease (KD) is an acute pediatric illness of unknown cause characterized by inflammation of the coronary arteries associated with fever and clinical criteria including rash, conjunctival injection, changes in lips or oropharyngeal mucosa, cervical lymphadenopathy, and changes in peripheral extremities^10^. Many of these clinical features overlap with MIS-C^11^. In response to the difficulty clinicians have in diagnosing and differentiating MIS-C and KD, we applied artificial intelligence (AI) to distinguish among children with MIS-C, KD, and other febrile illnesses characterized by similar clinical and laboratory features. Although there are reported differences such as older age, lower platelet count, and elevated inflammatory markers in MIS-C patients, none of these features alone was sufficient to diagnose MIS-C^4^. Currently, there is no published machine learning algorithm to differentiate between KD and MIS-C that has been externally validated.

We developed a two-stage AI model to classify patients as having MIS-C, KD, or other febrile illness using clinical signs and laboratory values that would be available at the time of a patient’s initial evaluation (**Figure 1**). Pediatric patients with other febrile illness, designated as febrile children (FC), were chosen based on the presence of fever and clinical or laboratory features suggestive of MIS-C and KD. The model in Stage 1 was trained to differentiate between MIS-C and other pediatric febrile conditions. The model in Stage 2 was trained to further classify patients falling into the “other” category as KD or other pediatric febrile illness. Since KD and MIS-C data distributions may vary across different sites, we incorporated a conformal prediction framework within the model^12^. Conformal prediction reduces false alarms by identifying unfamiliar samples in new patient populations when compared to the training cohort and assigns indeterminate labels rather than making spurious predictions. The laboratory tests incorporated into KIDMATCH are commonly obtained for pediatric patients in many outpatient and inpatient medical settings, and the clinical features would be easily assessable by frontline clinicians. The use of such readily available data enables KIDMATCH to be potentially deployable immediately across the United States without the need for specialized laboratory tests.

**Figure 1:**
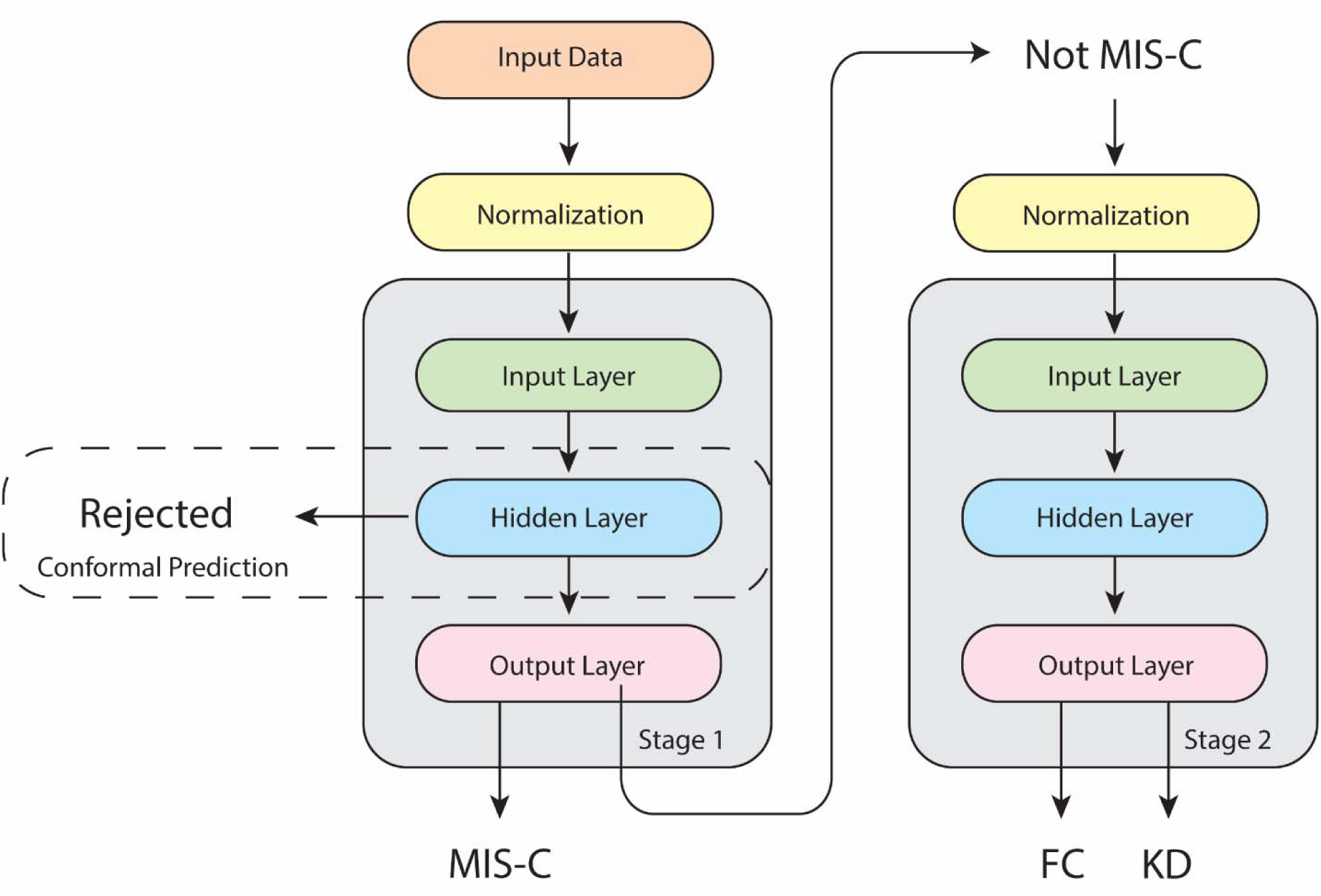
Model architecture. A patient could be classified as either MIS-C, febrile child (FC), or Kawasaki disease (KD) if the input data was not rejected by the conformal prediction framework.

## Methods

### Subjects

MIS-C patients were defined according to the CDC case definition^17^ and enrolled from May 14, 2020 to June 18, 2021. All MIS-C patients had positive antibody testing for either the nucleocapsid or spike protein of SARS-CoV-2 and none had received a SARS-CoV-2 vaccine. KD patients met the case definition of the American Heart Association^18^ for either complete or incomplete KD. In order to avoid the potential for misclassification, all KD subjects were enrolled from 2009-2019 prior to the pandemic. All KD subjects were diagnosed and treated by one of two highly experienced KD clinicians (J.C.B. and A.H.T.). FC subjects were also enrolled from 2009-2019 and met the following case definition: previously healthy child with fever for at least 3 days plus at least one of the clinical criteria for KD. Over 50% of the FC were referred for evaluation because of a clinical suspicion for KD. The final diagnoses for the FC were adjudicated 2-3 months after enrollment by two experienced pediatric clinicians who reviewed the clinical outcomes in the medical record and all available test results (**Supplemental Table 3**). A viral syndrome was defined as a self-limited illness that resolved without treatment and without apparent sequelae. Written consent or assent as appropriate was obtained from parents and subjects and the study was approved by the Institutional Review Boards (IRB) of the University of California San Diego (UCSD), Connecticut Children’s Medical Center, Children’s Hospital Los Angeles, Boston Children’s Hospital, and Children’s National Hospital. UCSD served as the central IRB of record for the CHARMS study participants.

### Data Preprocessing

We used age, the five classical clinical KD signs, and 17 laboratory measurements as features for KIDMATCH based on guidance from the clinician collaborators and availability of laboratory test results for the majority of the training cohort. The five clinical signs were rash, conjunctival injection, changes in lips or oropharyngeal mucosa, cervical lymphadenopathy, and changes in peripheral extremities. Laboratory data included white blood count, age-adjusted hemoglobin, platelets, polymorphonuclears or neutrophils, bands, lymphocytes, atypical lymphocytes, monocytes, eosinophils, absolute neutrophil count, absolute band count, erythrocyte sedimentation rate, C-reactive protein, alanine aminotransferase, gamma-glutamyl transferase, albumin, and sodium (**Table 1, Supplementary Table 2**). Due to the absence of bands and atypical lymphocytes in patients with automated differentials for the complete blood count, an indicator variable was added for the type of differential (0 = manual, 1 = automated). For samples with automated differentials, we imputed the values for bands, atypical lymphocytes, and absolute band counts using the mean of the respective feature. Outlier values defined as less than the 0.5^th^ percentile or greater than the 99.5^th^ percentile were set to the values of 0.5^th^ percentile if lower or 99.5^th^ percentile if higher. All other missing laboratory values were imputed using K-nearest neighbors as the mean of the respective feature for the 10 most similar samples from the training data. Data were normalized for each laboratory feature except hemoglobin by applying normalization transformations and then subtracting the mean and dividing by the standard deviation. Hemoglobin was normalized for age.

**Table 1:** Demographic and clinical characteristics of the patients used in internal validation from three clinical sites: Rady Children’s Hospital San Diego (n=1517), Connecticut Children’s Hospital (n=16), and Children’s Hospital of Los Angeles (n=50). Bolded characteristics are model features.

|  | MIS-C (n=131) | Kawasaki disease (n=775) | Febrile control (n=673) | p |
| --- | --- | --- | --- | --- |
| **Age, yrs\*** | 8.6 (4.3-11.3) | 2.9 (1.5-4.8) | 3.4 (1.5-5.9) | <0.0001 |
| Sex, male, n (%) | 77 (59) | 463 (59.7) | 394 (58.5) | NS |
| Ethnicity, n (%) |  |  |  |  |
| Asian | 5 (3.8) | 124 (16.0) | 47 (7.0) |  |
| African American | 14 (10.7) | 23 (3.0) | 10 (1.5) |  |
| White | 8 (6.1) | 167 (21.5) | 167 (24.8) | <0.0001 |
| Hispanic | 92 (70.2) | 275 (35.5) | 272 (40.4) |  |
| > 2 races or other | 12 (9.2) | 186 (24.0) | 114 (16.9) |  |
| No information | 0 | 0 | 63 (9.4) |  |
| Median Zmax† | 1.6 (0.8-2.4) | 1.7 (1.2-2.4) | NA | NS |
| Lowest LV Ejection Fraction, % | 57 (47-61) | 66 (63-70) | NA | <0.0001 |
| Illness day of Sample collection‡ | 4 (3-5.5) | 5 (4-7) | 5 (4-7) | NS |
| **Automated differential, n (%)** | 37 (28.2) | 33 (4.3) | 91 (13.5) | <0.0001 |
| Clinical Sign, n (%) |  |  |  |  |
| **Rash** | 74 (56.5) | 715 (92.3) | 442 (65.7) | <0.0001 |
| **Red Eyes** | 78 (59.5) | 725 (93.5) | 343 (51.0) | <0.0001 |
| **Oral Changes** | 48 (36.6) | 723 (93.3) | 304 (45.2) | <0.0001 |
| **Enlarged Cervical LN** | 22 (16.8) | 274 (35.4) | 151 (22.4) | <0.0001 |
| **Extremity Changes** | 18 (13.7) | 625 (80.6) | 131 (19.5) | <0.0001 |
| Laboratory data |  |  |  |  |
| **WBC, 10<sup>3</sup>/μL** | 9.8 (6.9-13.0) | 13.2 (10.4-17.0) | 9.5 (6.4-13.3) | <0.0001 |
| **Polys, %** | 71 (58-81) | 58 (47-69) | 47 (31-62) | <0.0001 |
| **Bands, %** | 12 (2-24) | 7 (2-15) | 5 (2-11) | <0.0001 |
| **Lymphocytes, %** | 11 (6-19) | 20 (12-31) | 32 (18-47) | <0.0001 |
| **Atypical lymphocytes, %** | 0 (0-1) | 0 (0-1) | 1 (0-3) | <0.0001 |
| **Monocytes, %** | 3 (1-5) | 6 (3-8) | 8 (5-11) | <0.0001 |
| **Eosinophils, %** | 1 (0-3) | 2 (1-4) | 0 (0-1.3) | <0.0001 |
| **ANC, /μL** | 7980 (5476-10525) | 8892 (6305-11768) | 4800 (2599-8208) | <0.0001 |
| **ABC, /μL** | 1160 (184-2168) | 891 (299-2095) | 438 (148-1205) | <0.0001 |
| **ALC, /μL** | 925 (549-1652) | 2565 (1412-3963) | 2730 (1633-4356) | <0.0001 |
| **ZHgb** | -1.7 (-3.0 to -0.8) | -1.3 (-2.3 to -0.5) | -0.3 (-1.3 to 0.5) | <0.0001 |
| **Platelet Count, $10^3/\text{mm}^3$** | 160 (112-222) | 339 (267-426) | 247 (184-322) | <0.0001 |
| **ESR, mm/h** | 49 (32-75) | 60 (39-75) | 29 (15-45) | <0.0001 |
| **CRP, mg/dL** | 18.9 (8.2-26.0) | 7.0 (4.3-16.9) | 2.9 (1.2-6.0) | <0.0001 |
| **ALT, IU/L** | 39 (22-63) | 46 (26-117) | 27 (19-38) | <0.0001 |
| **GGT, IU/L** | 36 (24-85) | 46 (18-128) | 15 (12-20) | <0.0001 |
| **Albumin, g/dL** | 3.6 (3.1-4.0) | 3.8 (3.5-4.2) | 4.1 (3.8-4.4) | <0.0001 |
| **Sodium, mmol/L** | 133 (130-135) | 137 (134-139) | 138 (136-139) | <0.0001 |

\*Values are median (Interquartile range (IQR)) unless otherwise specified; †: Maximum Z score (internal diameter normalized for body surface area) for the right and left anterior descending coronary arteries. ‡: Illness Day 1= first day of fever. | |: p-values were calculated by Mann-Whitney test for continuous variables between two groups, Kruskal-Wallis test for three groups and Chi-square test categorical variables. LN: lymph nodes, LV: left ventricle, WBC: white blood cell count, Polys: polymorphonuclears or neutrophils, ANC: absolute neutrophil count, ABC: absolute band count, ALC: absolute lymphocyte count, ZHgb: hemoglobin concentration normalized for age, PLT: platelet count, ESR: erythrocyte sedimentation rate, CRP: C-reactive protein, ALT: Alanine aminotransferase, GGT: gamma-glutamyltransferase, NS: not significant, NA: not applicable.

### Model Design

In KIDMATCH, we implemented a conformal prediction framework to reject out of distribution samples^12^ and separated the classification into two stages (**Figure 1**). If a test sample was rejected by the conformal prediction framework, no prediction was calculated. In Stage 1, the model calculated an MIS-C risk score between 0 and 1 for test samples with 1 being the highest MIS-C risk. In Stage 2, the model calculated a KD risk score between 0 and 1 where 1 is the highest risk for KD. Thresholds for each stage were set during internal validation to identify 95% of the MIS-C and KD cases in the test set for Stage 1 and Stage 2, respectively.

We trained a feedforward neural network on each of the stages using Tensorflow v.2.3.1 with a logistic regression using scikit-learn v0.24.2 as the baseline model. For both stages, the neural network was trained with the Adam optimizer at a learning rate of 0.01 and equally weighted batches of 100 and 200 samples from each class for Stage 1 and Stage 2, respectively. Each neural network consisted of an input layer (23 units), a single hidden layer (12 units, ReLu activation function, L2 regularization, 20% dropout rate), and a softmax output layer (2 units, binary cross entropy loss function).

### Model Training and Evaluation

We split the patients into training and test cohorts using an 80:20 split and employed a 10-fold cross validation to assess performance. Patients with any missing values were not considered for the test set. All patients were used from the training cohort in Stage 1 while MIS-C patients were omitted from the training cohort in Stage 2. Performance of the models were evaluated using accuracy, area under the receiver operating characteristic curve (AUC), positive predictive value (PPV), negative predictive value (NPV), false positive rate (FPR), and false negative rate (FNR) calculated at a minimum 95% sensitivity for the MIS-C and KD classifications for Stage 1 and Stage 2 respectively.

### Conformal Prediction

The trust sets used in the conformal prediction framework were constructed by filtering patients in the training cohort who had more than one missing value and MIS-C risk scores greater than the 95^th^ percentile (**Supplementary Table 1**). We then constructed a trust set for each of the three classifications (FC, KD, MIS-C) using patients from the training set. Finally, 200 randomly sampled patients were used for the FC and KD trust sets, and all MIS-C patients were used for the MIS-C trust set. We generated feature representations for the conformal prediction framework by passing the normalized features of the training samples through the first hidden layer of the neural network. Feature representations from the test samples were compared to each of the conformal trust sets using cosine similarity, and if a test sample was rejected by all three trust sets, then the model did not calculate risk scores for the test sample.

### Shapley Values

To explain the model predictions, we calculated the Shapley values for the test set using the SHAP Python library^13^. Normalized data from the training set was used as the background to compare normalized test set data for Stage 1 and Stage 2. 100 random background samples were used to calculate the Shapley values for each feature in the internal test set.

### Statistical Analysis

P-values were calculated by the Mann-Whitney test for continuous variables between two groups, the Kruskal-Wallis test for three groups, the Chi-square test for categorical variables within the cohort used for internal validation, and the DeLong test^19^ for AUC.

## Results

### Study population

A total of 1517 patients diagnosed from January 1, 2009 to October 1, 2021 with MISC (n=69), KD (n=775), or other febrile illnesses (n=673) were identified from a REDCap database at the Kawasaki Disease Research Center in the University of California San Diego (UCSD). Laboratory tests and clinical signs were obtained at the time of initial evaluation and prior to treatment for all patients. To improve the generalizability of the model, we added MIS-C patients from Connecticut Children’s Hospital (n=16) and Children’s Hospital of Los Angeles (n=50) when training the model for a total of 135 MIS-C patients during internal validation (**Table 1**). External validation was performed using MIS-C clinical data (n=175) enrolled from May 14, 2020 to June 18, 2021 from the following sites: Boston Children’s Hospital (n=50), Children’s nc Hospital (n=42), and the CHARMS consortium (n=83), a 14-site multicenter database of MIS-C patients funded by the Patient Centered Outcomes Research Institute (PCORI) and housed at UCSD. In a comparison of laboratory data among the groups, MIS-C patients had higher band counts, lower sodium levels, lower platelet counts, and higher C-reactive protein (p < 0.0001) compared to the FC and KD cohorts, consistent with prior reports^1,4^ (**Table 1**).

### Internal Validation

The 10-fold cross validation results for Stage 1 and 2 are shown in **Table 2**. The neural network in Stage 1 had similar accuracy, area under the receiver operating characteristic curve (AUC), positive predictive value (PPV), negative predictive value (NPV), false positive rate (FPR), and false negative rate (FNR) compared to the logistic regression baseline in the validation set when classifying samples as MIS-C or not MIS-C. In Stage 2, the neural network compared favorably to the logistic regression baseline in terms of accuracy, PPV, and FPR when classifying samples as FC or KD based on thresholds set at 95% sensitivity for KD samples (**Table 2**). We chose 95% sensitivity based on clinician feedback to avoid missing true positive subjects.

**Table 2:**
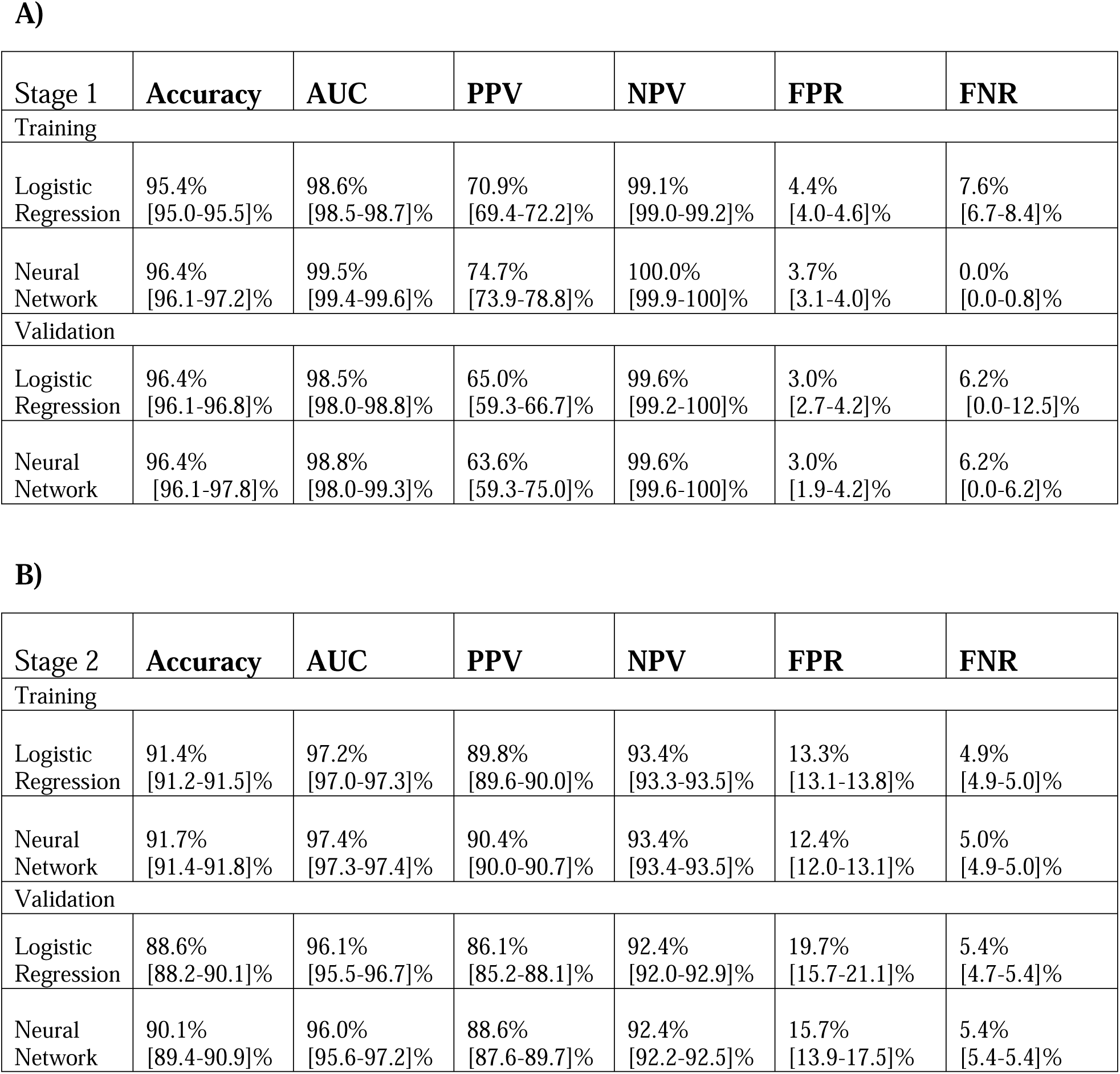
10-fold cross validation performance metrics for the training and validation cohorts in (A) Stage 1 and (B) Stage 2. All values reported as median [IQR].

| Stage 1 | Accuracy | AUC | PPV | NPV | FPR | FNR |
| --- | --- | --- | --- | --- | --- | --- |
| Training |  |  |  |  |  |  |
| Logistic Regression | 95.4%<br>[95.0-95.5]% | 98.6%<br>[98.5-98.7]% | 70.9%<br>[69.4-72.2]% | 99.1%<br>[99.0-99.2]% | 4.4%<br>[4.0-4.6]% | 7.6%<br>[6.7-8.4]% |
| Neural Network | 96.4%<br>[96.1-97.2]% | 99.5%<br>[99.4-99.6]% | 74.7%<br>[73.9-78.8]% | 100.0%<br>[99.9-100]% | 3.7%<br>[3.1-4.0]% | 0.0%<br>[0.0-0.8]% |
| Validation |  |  |  |  |  |  |
| Logistic Regression | 96.4%<br>[96.1-96.8]% | 98.5%<br>[98.0-98.8]% | 65.0%<br>[59.3-66.7]% | 99.6%<br>[99.2-100]% | 3.0%<br>[2.7-4.2]% | 6.2%<br>[0.0-12.5]% |
| Neural Network | 96.4%<br>[96.1-97.8]% | 98.8%<br>[98.0-99.3]% | 63.6%<br>[59.3-75.0]% | 99.6%<br>[99.6-100]% | 3.0%<br>[1.9-4.2]% | 6.2%<br>[0.0-6.2]% |

**B)**
| Stage 2 | Accuracy | AUC | PPV | NPV | FPR | FNR |
| --- | --- | --- | --- | --- | --- | --- |
| Training |  |  |  |  |  |  |
| Logistic Regression | 91.4%<br>[91.2-91.5]% | 97.2%<br>[97.0-97.3]% | 89.8%<br>[89.6-90.0]% | 93.4%<br>[93.3-93.5]% | 13.3%<br>[13.1-13.8]% | 4.9%<br>[4.9-5.0]% |
| Neural Network | 91.7%<br>[91.4-91.8]% | 97.4%<br>[97.3-97.4]% | 90.4%<br>[90.0-90.7]% | 93.4%<br>[93.4-93.5]% | 12.4%<br>[12.0-13.1]% | 5.0%<br>[4.9-5.0]% |
| Validation |  |  |  |  |  |  |
| Logistic Regression | 88.6%<br>[88.2-90.1]% | 96.1%<br>[95.5-96.7]% | 86.1%<br>[85.2-88.1]% | 92.4%<br>[92.0-92.9]% | 19.7%<br>[15.7-21.1]% | 5.4%<br>[4.7-5.4]% |
| Neural Network | 90.1%<br>[89.4-90.9]% | 96.0%<br>[95.6-97.2]% | 88.6%<br>[87.6-89.7]% | 92.4%<br>[92.2-92.5]% | 15.7%<br>[13.9-17.5]% | 5.4%<br>[5.4-5.4]% |

We selected the model with the highest accuracy during the 10-fold cross-validation at each stage to use in the final model. To ensure model generalizability and similar performance across external sites, we constructed a conformal prediction framework using the training samples from the final model^12^. Briefly, we selected the parsimonious combination of missingness and risk score with the highest weighted F1 score to construct the conformal trust sets (**Supplementary Figure 2, Supplementary Table 1**). This approach rejected 3/149 (2%) of the FC samples, 6/165 (3.6%) of the KD samples, and none of the MIS-C samples in the validation set. The ROC curves for the final model indicated that the neural networks had high sensitivity and specificity with an AUC of 0.98 in Stage 1 and 0.97 in Stage 2 (**Figure 2**).

**Figure 2:**
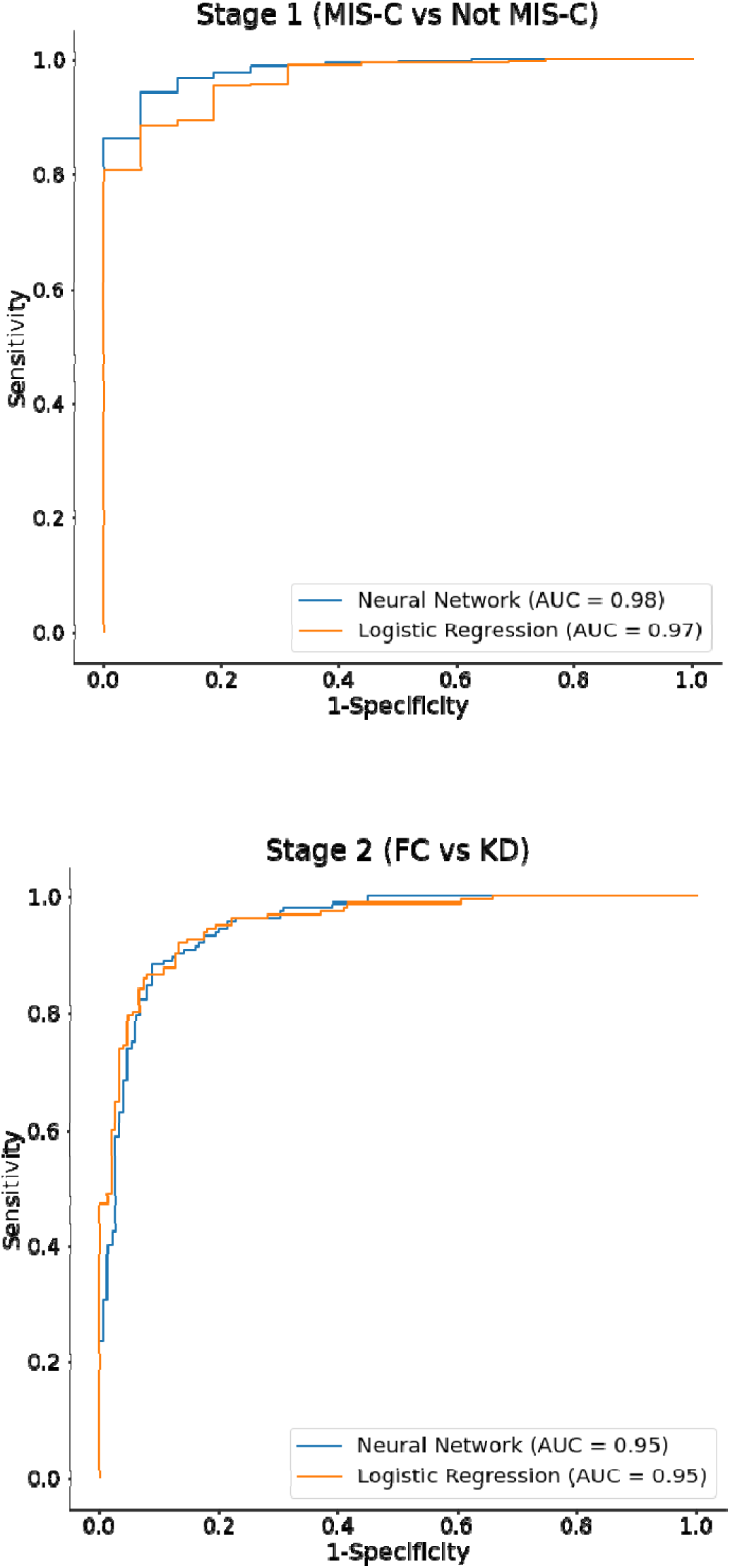
Receiver operating curves for the best cross-validation fold of (A) Stage 1 and (B) Stage 2 in terms of AUC.

The neural networks trained for each stage in the final model showed robust performance when setting thresholds (Stage 1 – 0.36, Stage 2 – 0.60) at a 95% sensitivity level (**Figure 2**). Although there was no statistical difference in the AUC for Stage 1 (p = 0.174) and Stage 2 (p = 0.594), the neural networks were chosen for the final model as the conformal prediction framework relied on feature representations that could not be calculated using logistic regression. In addition, the majority of patients at UCSD had a complete blood count with manual differential, but the external sites had a significant proportion of automated differentials. The neural networks were able to adjust for the difference between manual complete blood counts and automated differential counts effectively by incorporating an indicator variable as input to the model.

Devising a diagnostic test for two diseases for which there is no gold standard test presents special challenges. As a sensitivity analysis, we tested KD patients with coronary artery aneurysms and MIS-C patients who had a reduced left ventricular ejection fraction as characteristic patient subsets who were unlikely to be misclassified by clinicians. The final model correctly assigned 94% of the 164 KD patients with coronary artery aneurysms and 95% of the MIS-C patients with reduced ejection fraction (**Supplementary Figure 1**).

### Feature Importance

To determine how the features contributed to the model predictions, we used Shapley values, specifically the Shapley Additive exPlanations (SHAP) method^13^. In Stage 1, the most important features that distinguished MIS-C vs non MIS-C patients were serum sodium, platelet count, neutrophils, and C-reactive protein (**Figure 3**). The patterns observed were consistent with published reports of the laboratory testing characteristics of MIS-C patients^1,4^. In Stage 2, changes in peripheral extremities, erythrocyte sedimentation rate, conjunctival injection, and erythema of the lips or oropharyngeal mucosa were the most important features for differentiating between FC and KD. Three of the four aforementioned features are clinical signs used by clinicians to diagnose KD, so it is not surprising that the presence of one or more of these clinical signs contributed to a higher Stage 2 risk score and higher probability of KD. The next most important features were age, with younger patients more likely to have KD, and gamma glutamyl transferase (GGT), for which higher levels indicate hepatobiliary inflammation often observed in KD^14^.

**Figure 3:**
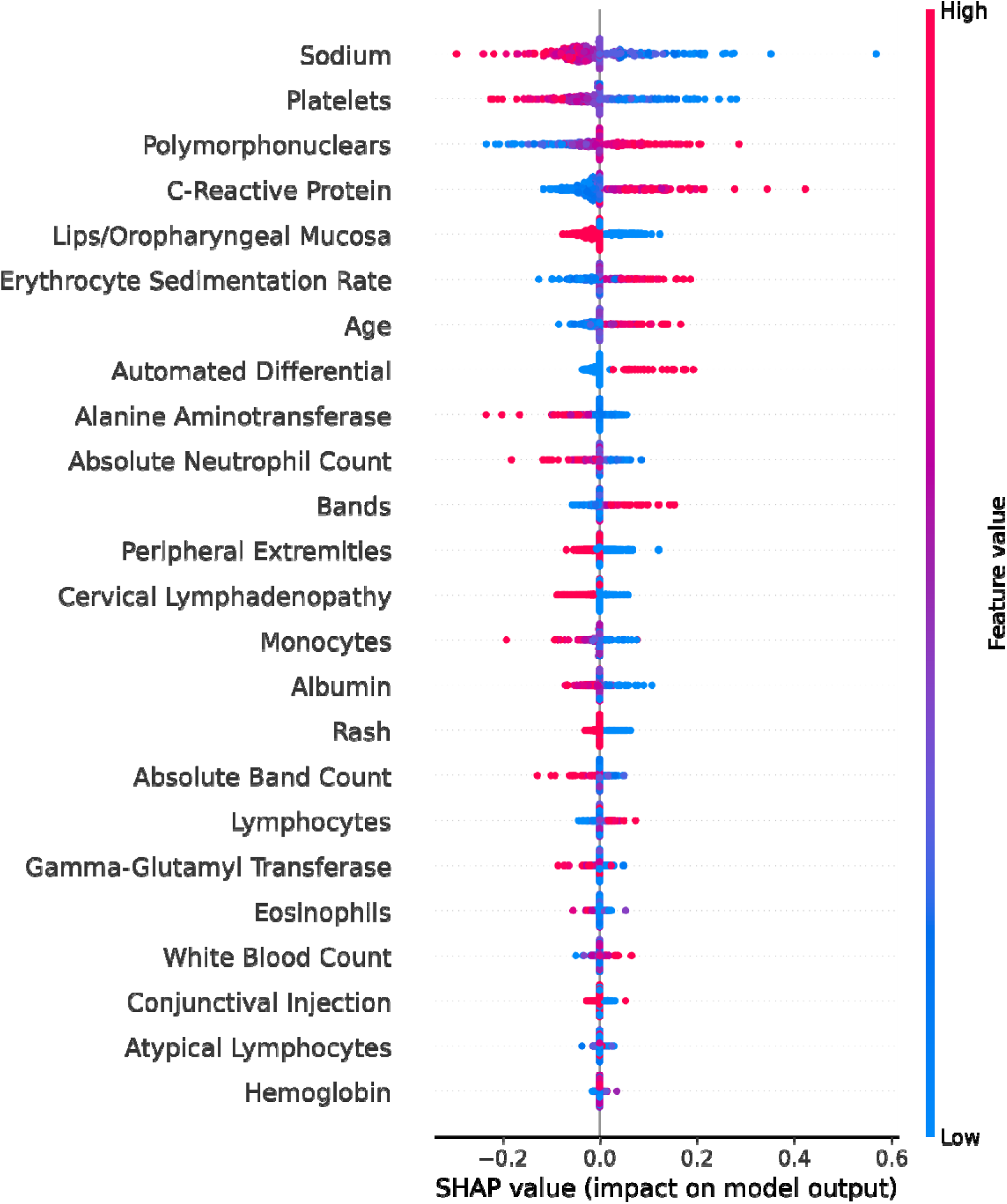
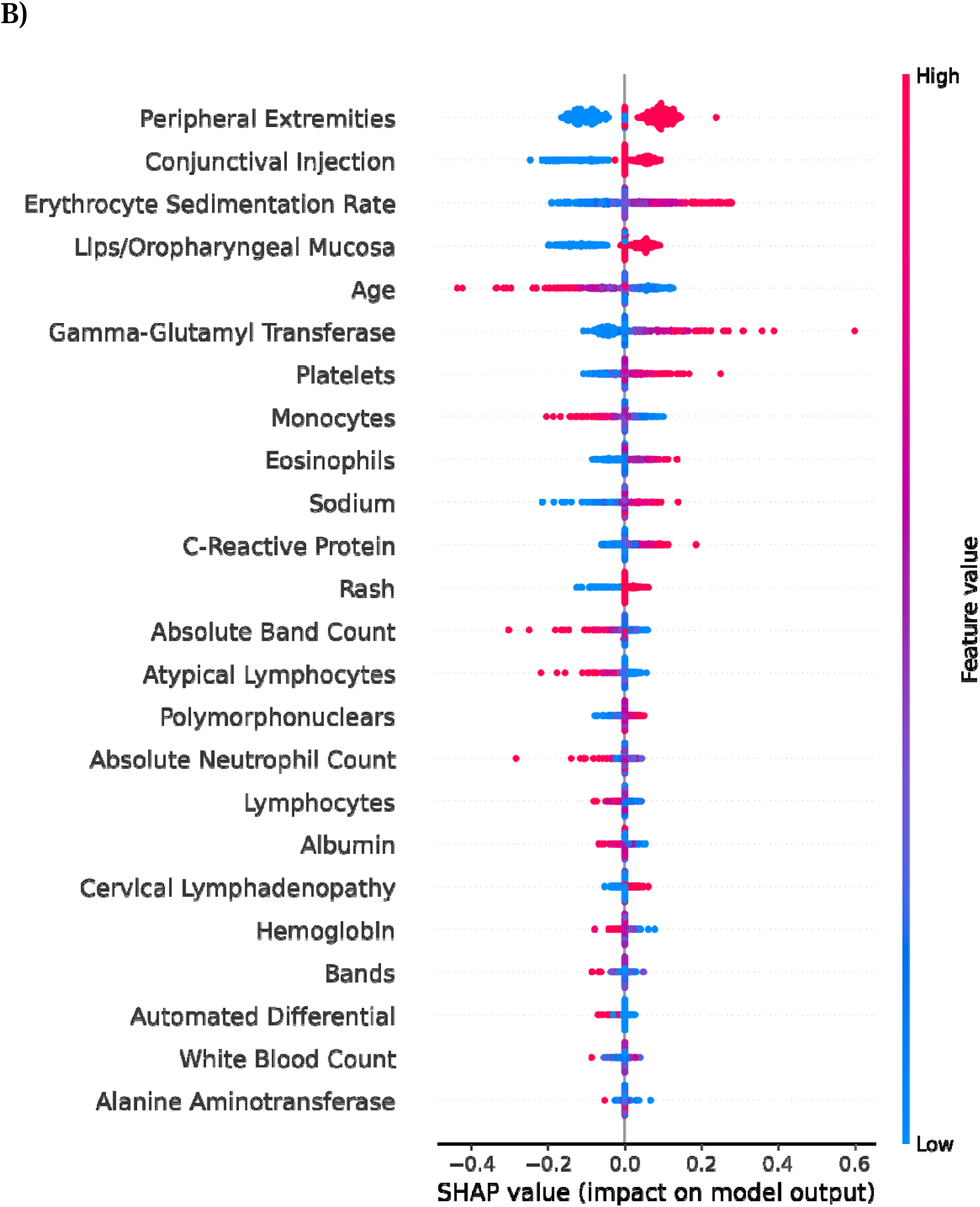
SHAP summary plot for (A) Stage 1 and (B) Stage 2 with raw feature values. A positive SHAP value means the corresponding feature value increases the risk score and vice versa. In Stage 1, a higher risk score indicates a higher probability of MIS-C. In Stage 2, a higher risk score indicates a higher probability of KD. Features are ranked in order of importance from top to bottom.

### External Validation

We externally validated KIDMATCH using MIS-C patients from the CHARMS consortium, Boston Children’s Hospital (Boston), and Children’s National Hospital (CNH). Our conformal prediction framework rejected 2/83 (2.4%), 1/50 (2%), and 2/42 (4.8%) samples from the CHARMS consortium, Boston, and CNH respectively. KIDMATCH accurately predicted MIS-C in 93.8% of CHARMS patients, 95.9% of Boston patients, and 90% of CNH patients after conformal prediction (**Table 3**). Examination of the Stage 1 risk scores for each site revealed most MIS-C patients were confidently classified as MIS-C (risk score > 0.8) (**Figure 4**).

**Table 3:** Predicted classifications of MIS-C patients from external sites. Percentages are based on the total number of patients from each site that were not rejected by conformal prediction.

| Site | MIS-C Subjects Predicted As |  |  |  |
| --- | --- | --- | --- | --- |
|  | FC | KD | MIS-C | Total |
| CHARMS | 3 (3.7%) | 2 (2.5%) | 76 (93.8%) | 81 |
| Boston Children's Hospital | 2 (4.1%) | 0 (0%) | 47 (95.9%) | 49 |
| Children's National Hospital | 3 (7.5%) | 1 (2.5%) | 36 (90%) | 40 |

**Figure 4:**
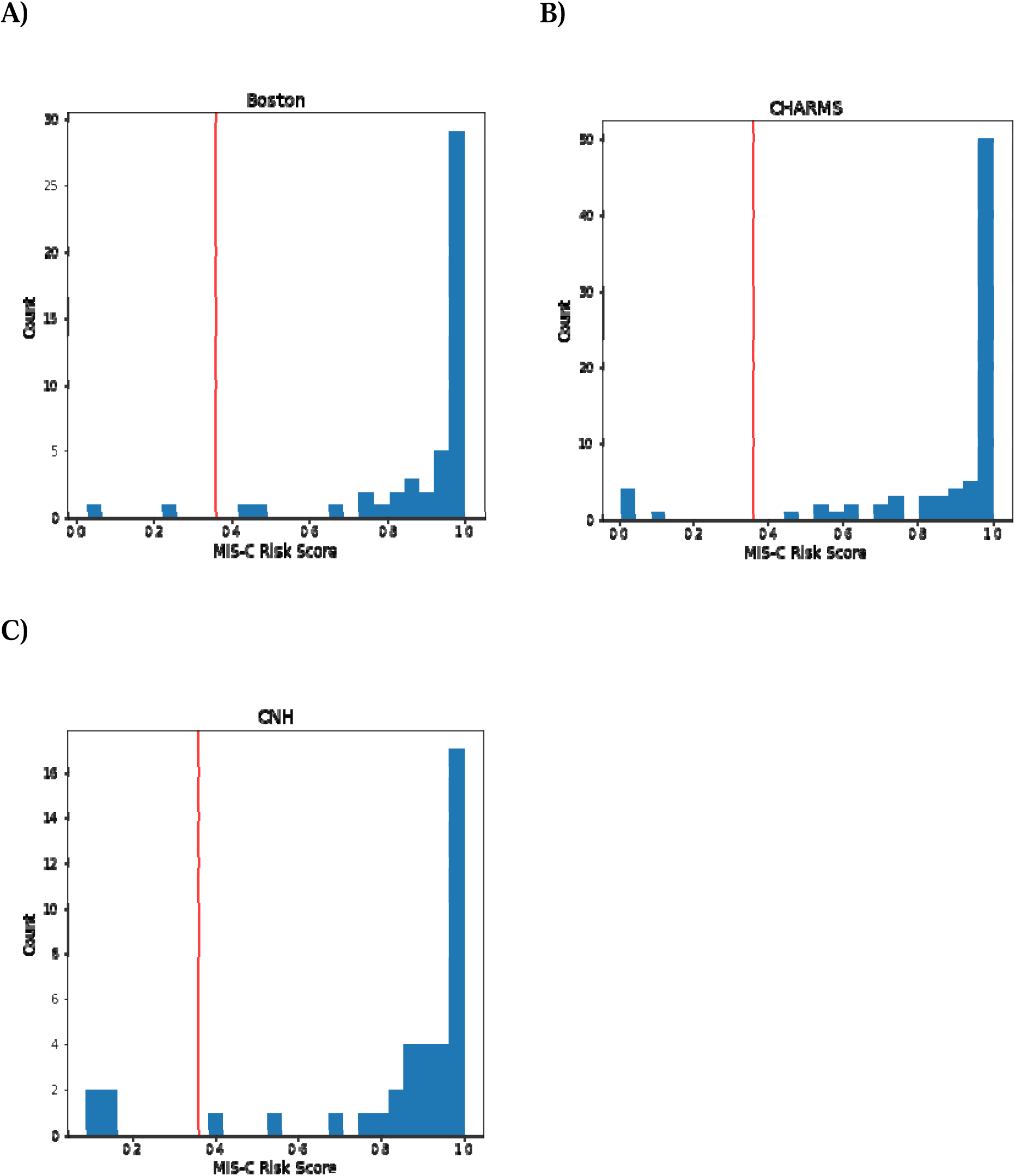
Stage 1 MIS-C external validation risk scores from (A) Boston, (B) CHARMS, and (C) CNH patients. Patients with risk scores greater than the threshold (0.36, denoted by a red line) were classified as MIS-C, and patients with lower risk scores were passed to Stage 2.

Although we had only 26 MIS-C patients in the internal validation testing cohort, KIDMATCH generalized well to external MIS-C cohorts with 90% or greater accuracy at all three external sources despite missingness of 1-4 features, most often GGT. Interestingly, the predictive performance was not replicated consistently across all sites. CNH had the lowest prediction accuracy at 90%. Further investigation revealed that their laboratory values for albumin were significantly lower than the MIS-C training distribution (2.9 [2.5-3.2] vs. 3.6 [3.2-4.0] g/dL, median [IQR], p < 0.001) due to differences in the test platform used by that clinical laboratory (**Supplementary Figure 3**). In addition, all misclassified MIS-C patients from CNH had a normal serum sodium of 138 mmol/L or higher, and the distribution of serum sodium values from this laboratory was significantly higher compared to the other MIS-C clinical sites (136 [134-139] vs. 133 [130-135], median [IQR], p < 0.01). While these values deviated from those observed in other sites, we cannot dismiss the possibility that the serum albumin and sodium values are valid as outlier values have been observed in MIS-C patients within the training cohort. Fortunately, the model showed consistent performance when handling samples with outlier values. The reliance of the Stage 1 algorithm on characteristic MIS-C laboratory test values such as low serum sodium and low platelet count increases the probability of misclassification when presented with normal values from a MIS-C patient (**Supplementary Figure 4**). However, the model enables clinicians to explore how the relevant features are contributing to the risk score and adjust their clinical judgement accordingly.

## Discussion

We present a machine learning model for screening patients with MIS-C, KD, or similar febrile illnesses using clinical signs and laboratory data routinely collected during the initial evaluation of these patients. To date, this is the first application of artificial intelligence to aid in the diagnosis of MIS-C and differentiate it from KD. KIDMATCH has the ability to reject test samples that are outside of the distribution in the training set, which provides a measure of confidence by statistically identifying outlier inputs. It is interpretable on a case-by-case basis by examining the most important features and whether they impact the MIS-C risk score positively or negatively. KIDMATCH showed consistent performance across different hospitals and the conformal framework identified outlier samples that would have been misclassified otherwise, thus demonstrating our algorithm can be applied in diverse clinical settings. A web-based user interface was developed using *Streamlit*, an open-source framework for building applications in Python, to assist clinicians with calculation of the proposed risk scores and assessment of the top factors contributing to risk (**Supplementary Figure 5**).

A strength of our work is the universal availability of the required features in the majority of healthcare settings and the validation using external cohorts. The only published model addressing a machine learning approach to MIS-C diagnosis includes features that may not be available in many clinical settings such as ferritin and D-dimer^15^. In addition, the model was not tested against febrile pediatric control patients and was not externally validated.

We recognize limitations to our work due to the limited availability of FC and KD data for external validation. There is no gold standard for KD or MIS-C diagnosis. Thus, we cannot exclude some degree of misdiagnosis in either the training or test set. However, the validation performance demonstrated internal consistency with the signs and laboratory tests used by KIDMATCH. It is unknown how the model would perform on FC and KD patients from other hospitals as we were unable to obtain these data. The thresholds established during internal validation may not be generalizable to different sites and shifting the threshold may be required to adjust for different prevalence rates. However, the high model AUC means that the model can be used to effectively prioritize febrile patients for further evaluation of MIS-C or KD. A key step for deployment will be to establish standardized conditions for use so the algorithm is applied to the appropriate patients. The current algorithm is only optimized for laboratory test values collected at the time of initial evaluation, and it is unknown how it would perform with data collected at a later timepoint. It is also unknown how end users should deal with patients flagged as indeterminate, but a possible solution could be to order more specialized tests such as ferritin, troponin, BNP/NT-proBNP, and D-dimer as well as IgG antibody to SARS-CoV-2 as is routine practice for suspected MIS-C patients^16^.

Based on these data, the proposed algorithm is a generalizable and trustworthy tool for the diagnosis of MIS-C and KD during the initial evaluation of the patient. Future work includes retrospective validation on external FC and KD patients and prospective validation on MIS-C patients as well as implementing KIDMATCH within the clinical workflow. As the first externally validated machine learning solution for the diagnosis of MIS-C, KIDMATCH has the potential to aid frontline clinicians and improve patient outcomes through timely diagnosis.

## Data Availability

De-identified data has been compiled from multiple sites across the United States. Requests for data will require approval from UCSD and partner institutions independently.

## Author Contributions

J.Y.L., S.H., X.B.L., H.J.C., S.N., and J.C.B. were involved in the original conception of the work. S.C.R., C.S., N.S., A.H.T., M.A.G., J.T.K., A.H.H., J.C.S., S.M., J.R.S., S.M., A.D., J.W.N., E.A., R.L.D., and members of the Pediatric Emergency Medicine Kawasaki Disease Research Group and the CHARMS Study Group assisted with patient enrollment and data acquisition. J.Y.L. and S.N. developed the model architectures, conducted the experiments, and analyzed the data. J.Y.L., S.N., and J.C.B. wrote the initial draft of the manuscript. All other authors contributed feedback and approved the final draft.

## Acknowledgements

This work was supported by NIH grants (R61HD105590 to J.C.B, R01HL140898 to J.C.B. and A.H.T) and a grant from the Patient-Centered Outcomes Research Institute (CER-1602-3447 to J.C.B). J.Y.L. was supported by a predoctoral fellowship from the National Library of Medicine (T15LM011271). We would like to thank Supreeth Shashikumar, PhD, for technical advice and support with the conformal prediction framework.

## Contributor Information

### The Pediatric Emergency Medicine Kawasaki Disease Research Group

Naomi Abe, Lukas Austin-Page, Amy Bryl, J. Joelle Donofrio-Ödmann, Atim Ekpenyong, Michael Gardiner, David J. Gutglass, Margaret B. Nguyen, Kristy Schwartz, Stacey Ulrich Tatyana Vayngortin, Elise Zimmerman

### The CHARMS Study Group

Marsha Anderson, Jocelyn Y. Ang, Emily Ansusinha, Negar Ashouri, Joseph Bocchini, Laura D’Addese, Roberta DeBiasi, Samuel Dominguez, Maria Pila Gutierrez, Ashraf S. Harahsheh, Michelle Hite, Pei-Ni Jone, Madan Kumar, John J. Manaloor, Marian Melish, Lerraughn Morgan, JoAnne E. Natale, Allison Rometo, Margalit Rosenkranz, Anne H. Rowley, Nichole Samuy, Paul Scalici, Michelle Sykes

Marsha Anderson - Children’s Hospital Colorado, Aurora, CO, USA

Jocelyn Y. Ang - Children’s Hospital of Michigan, Detroit, MI, USA, Wayne State University School of Medicine, Detroit, MI, USA, Central Michigan University College of Medicine, Mt. Pleasant, MI, USA

Emily Ansusinha - Children’s National Hospital, Washington, DC, USA

Negar Ashouri - CHOC Children’s Hospital, Orange, CA, USA

Joseph Bocchini - Knighton Children’s Health - Tulane University, Shreveport, LA, USA

Laura D’Addese - Joe Dimaggio Children’s Hospital, Hollywood, FL, USA

Roberta DeBiasi - Children’s National Hospital, Washington, DC, USA, George Washington University School of Medicine & Health Sciences, Washington, DC, USA.

Samuel Dominguez - Children’s Hospital Colorado, Aurora, CO, USA

Maria Pila Gutierrez - Joe Dimaggio Children’s Hospital, Hollywood, FL, USA

Ashraf S. Harahsheh - Children’s National Hospital, Washington, DC, USA, George Washington University School of Medicine & Health Sciences, Washington, DC, USA.

Michelle Hite - Children’s Hospital Colorado, Aurora, CO, USA

Pei-Ni Jone - Children’s Hospital Colorado, Aurora, CO, USA

Madan Kumar - The University of Chicago Department of Pediatrics, Chicago, IL, USA

John J. Manaloor - Indiana University School of Medicine, Indianapolis, IN, USA

Marian Melish - University of Hawaii, Kapi’olani Medical Specialists, Honolulu, HI, USA

Lerraughn Morgan - Valley Children’s Healthcare, Madera, CA, USA

JoAnne E. Natale - UC Davis Children’s Hospital, Sacramento, CA, USA

Allison Rometo - UPMC Children’s Hospital of Pittsburgh, Pittsburgh, PA, USA

Margalit Rosenkranz - UPMC Children’s Hospital of Pittsburgh, Pittsburgh, PA, USA

Anne H. Rowley - The Ann & Robert H. Lurie Children’s Hospital of Chicago, Chicago, IL, USA

Nichole Samuy - University of Alabama at Birmingham, Birmingham, AL, USA

Paul Scalici - University of Alabama at Birmingham, Birmingham, AL, USA

Michelle Sykes - Valley Children’s Healthcare, Madera, CA, USA

## Supplementary Information

**Supplementary Figure 1:**
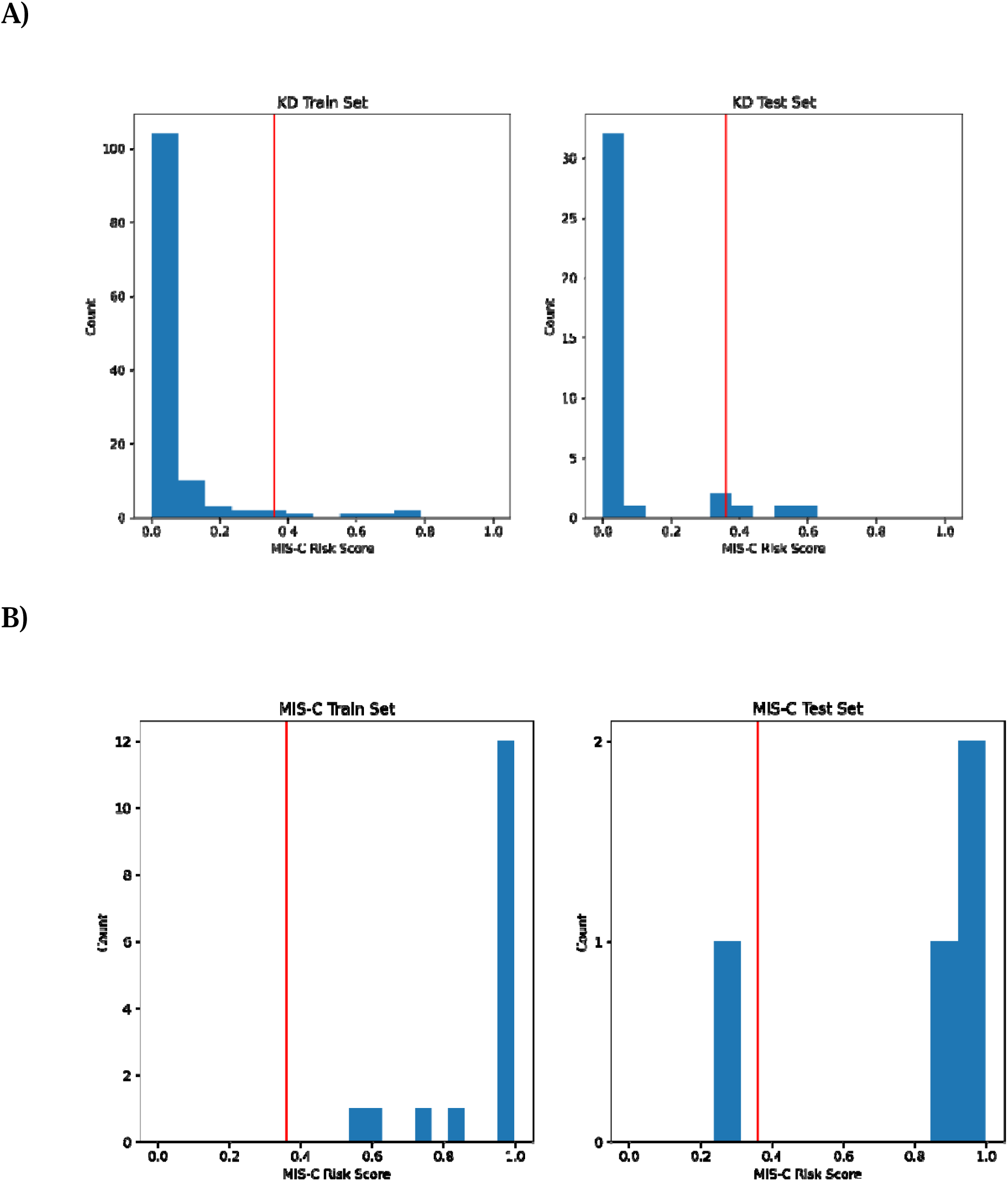
Stage 1 MIS-C risk scores for UCSD (A) KD patients with aneurysms (n=164) and (B) MIS-C patients with reduced ejection fraction (n=20) in the training and test set. Coronary artery aneurysms were defined as a maximum Z score (internal diameter normalized for body surface area) for the right and left anterior descending coronary arteries > 2.5. Reduced ejection fraction in MIS-C patients was defined as a left ventricular ejection fraction <55%. Patients with risk scores greater than the threshold (0.36, denoted by a red line) were classified as MIS-C. In the KD cohort, 120/126 patients in the training set and 34/38 individuals in the test set were classified correctly as Not MIS-C. In the MIS-C cohort, 16/16 individuals in the training set and 3/4 individuals in the test set were classified correctly as MIS-C.

**Supplementary Figure 2:**
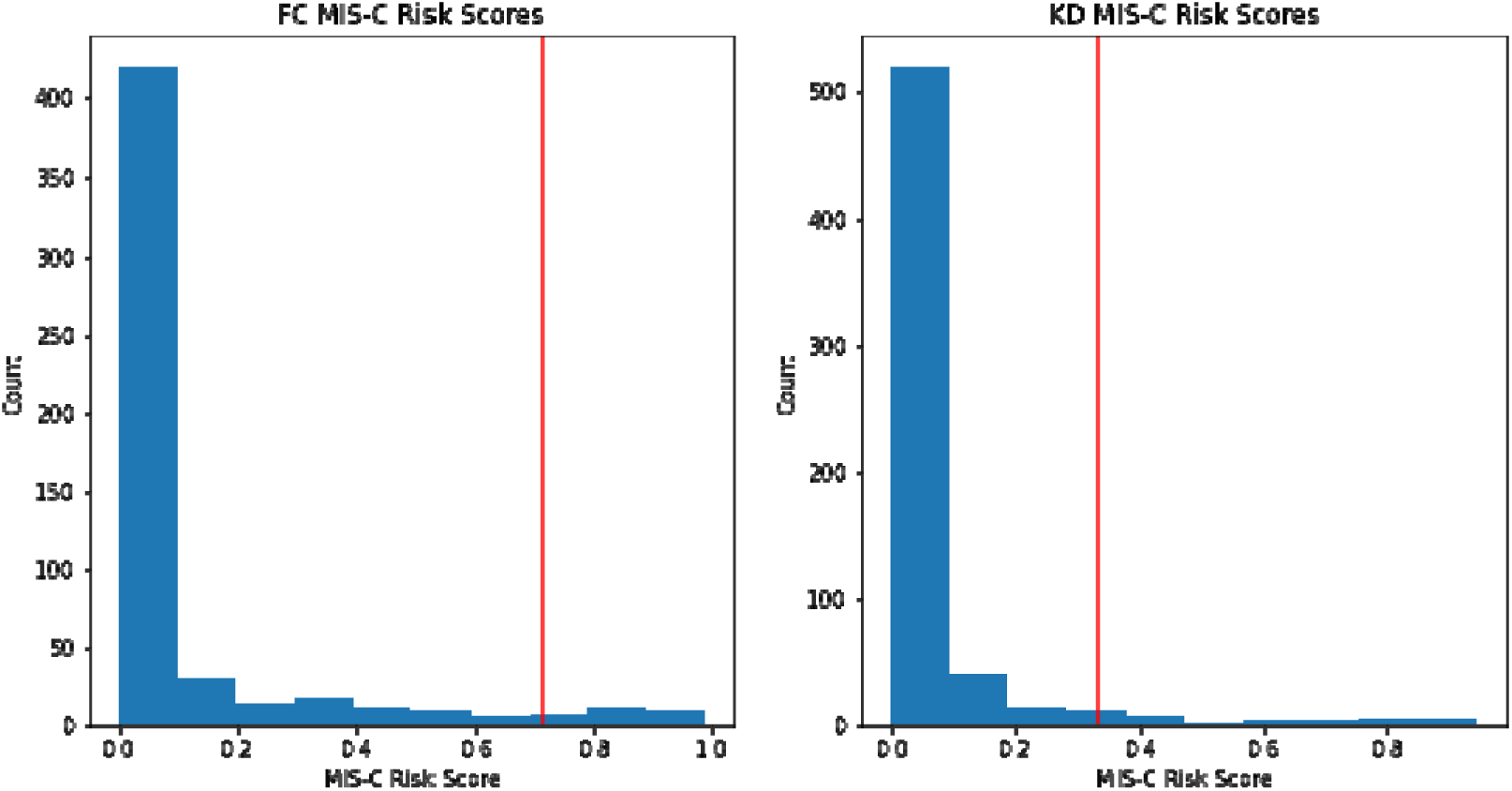
MIS-C risk scores for the FC and KD patients in the training cohort with at maximum 1 missing value. The 95^th^ percentile for each cohort is denoted by a vertical red line. Random samples were taken from the group of FC and KD patients with MIS-C risk scores below the 95^th^ percentile to create the conformal trust sets.

**Supplementary Figure 3:**
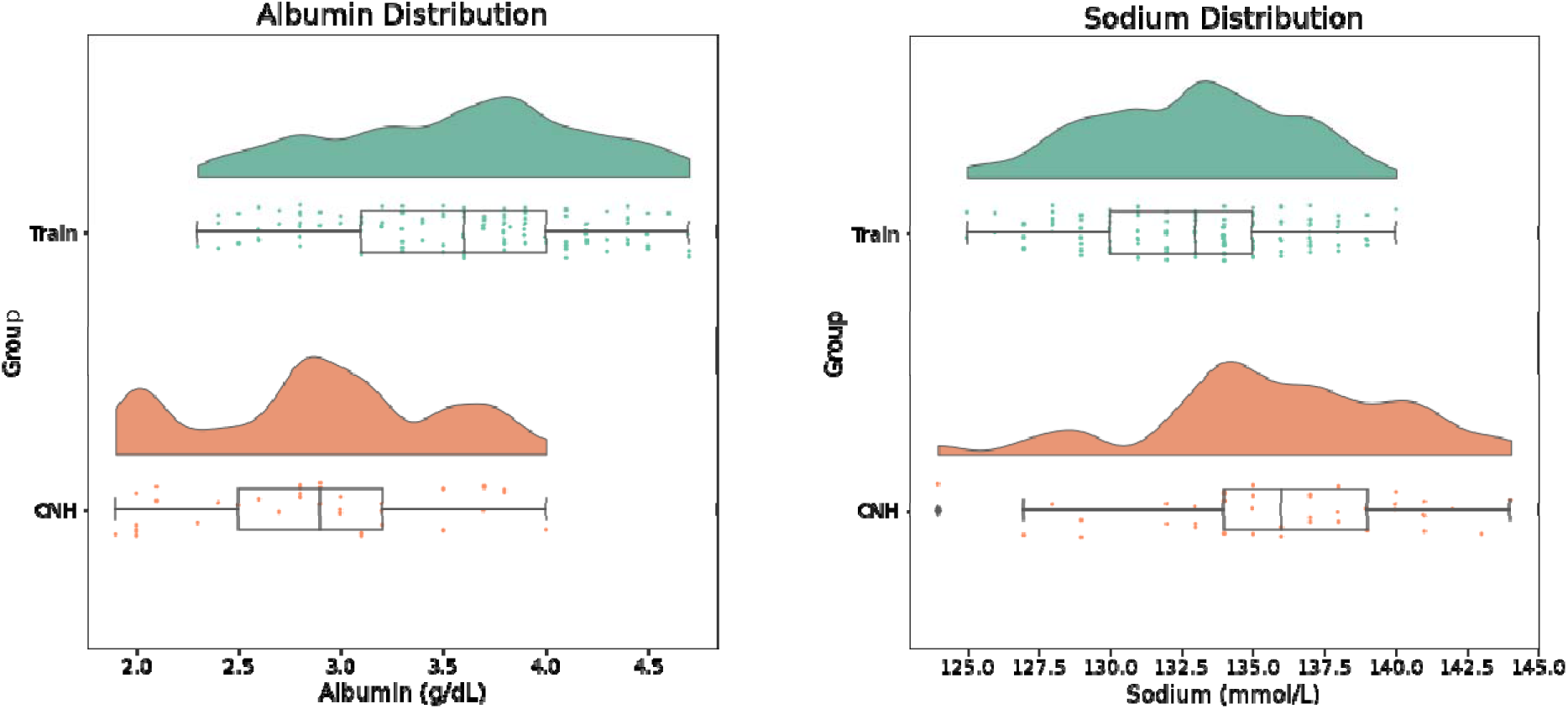
(A) Albumin and (B) sodium distributions from MIS-C patients in the training set (Train) compared to Children’s National Hospital (CNH).

**Supplementary Figure 4:**
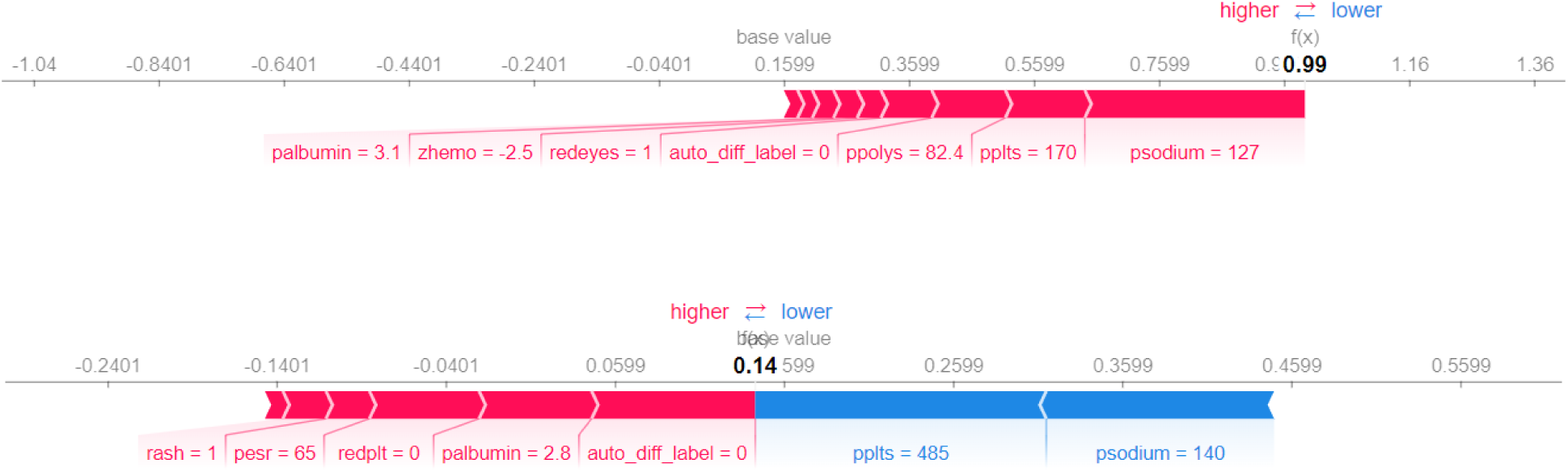
SHAP force plots from a (A) correctly classified MIS-C patient (MIS-C risk score of 0.99) and (B) incorrectly classified MIS-C patient (MIS-C risk score of 0.l4). Red indicates a feature has a positive contribution to the risk score and blue indicates the opposite. Values for the most important features to each patient’s risk score are listed at the bottom of the plot.

**Supplementary Figure 5:**
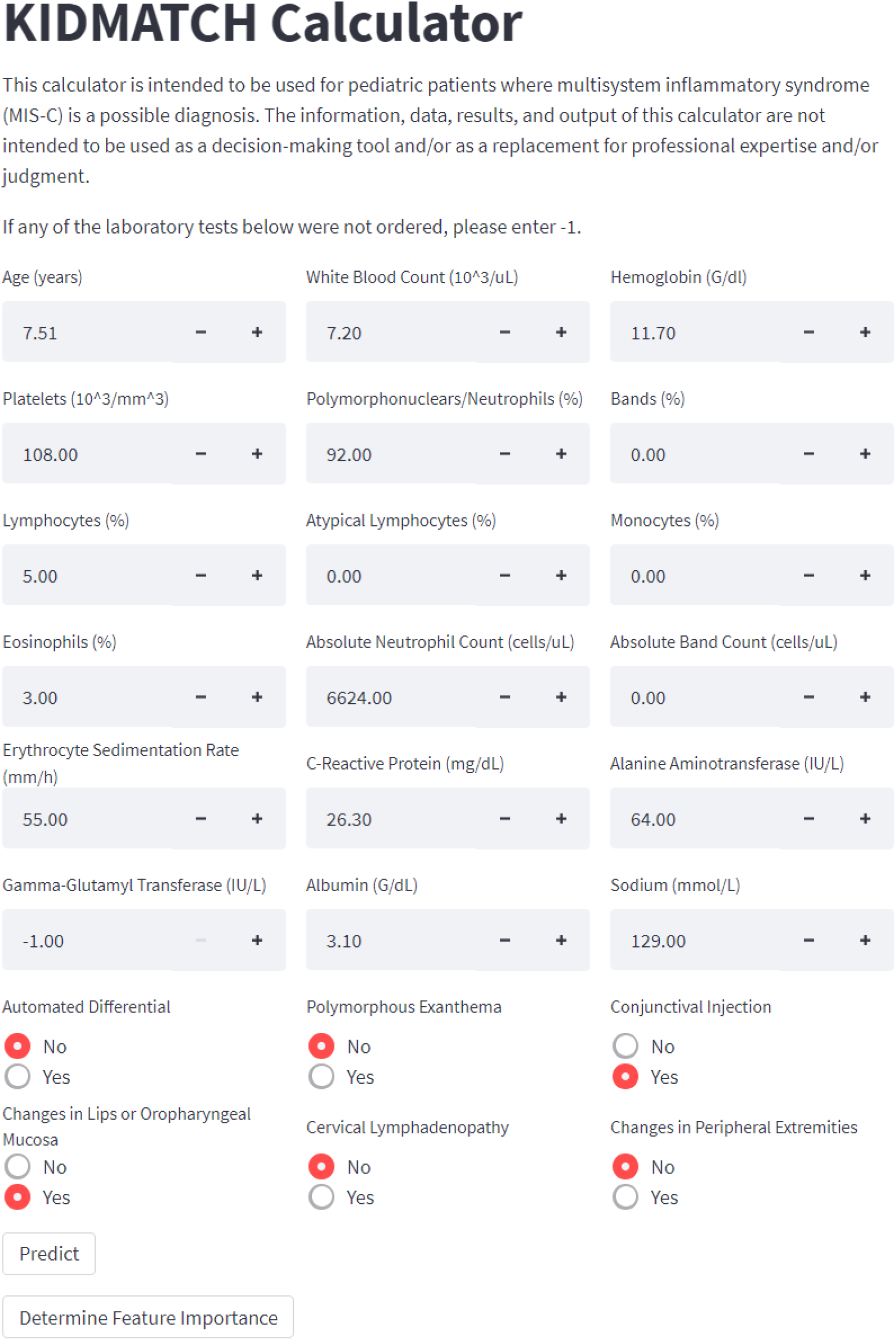
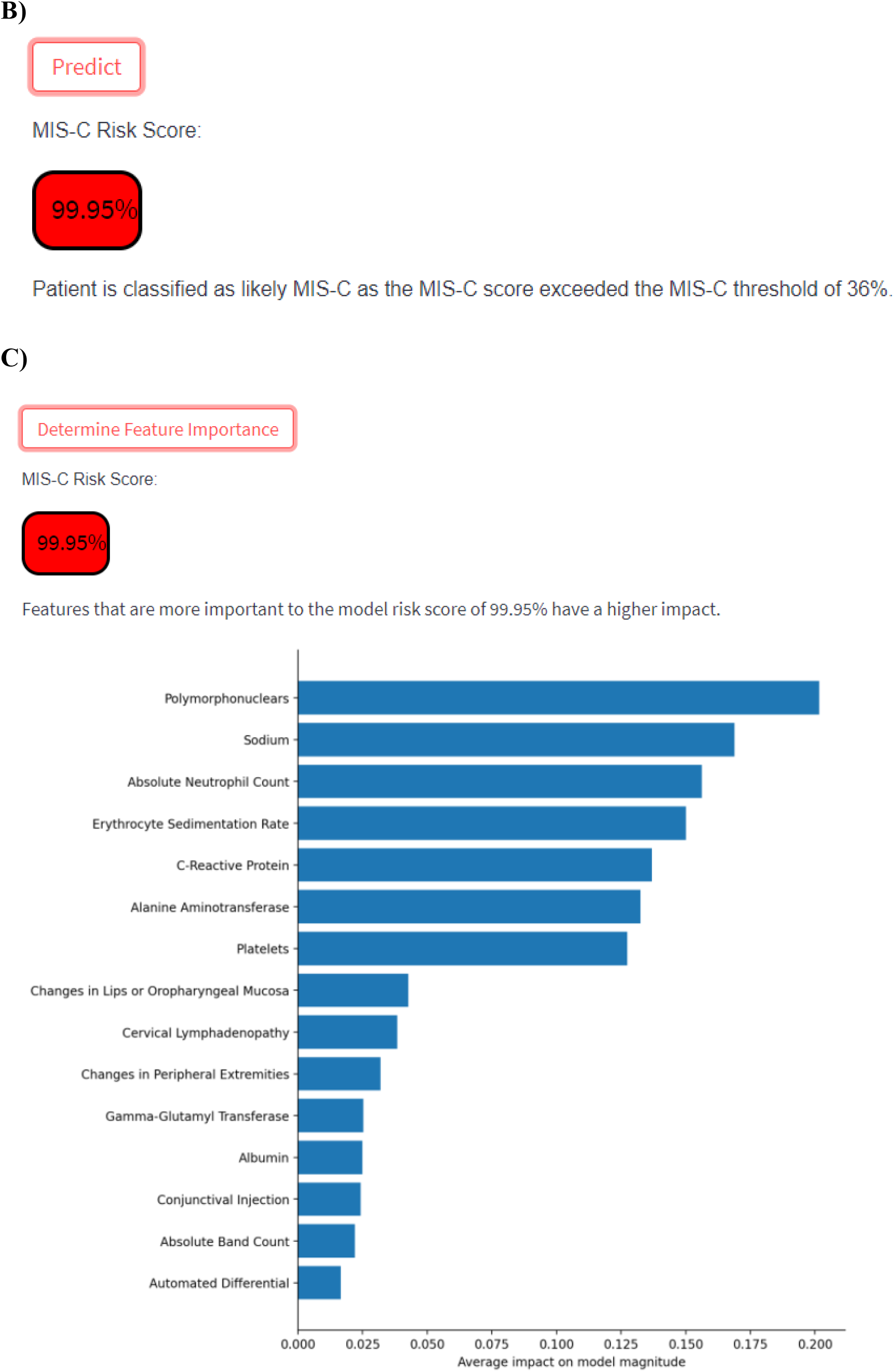
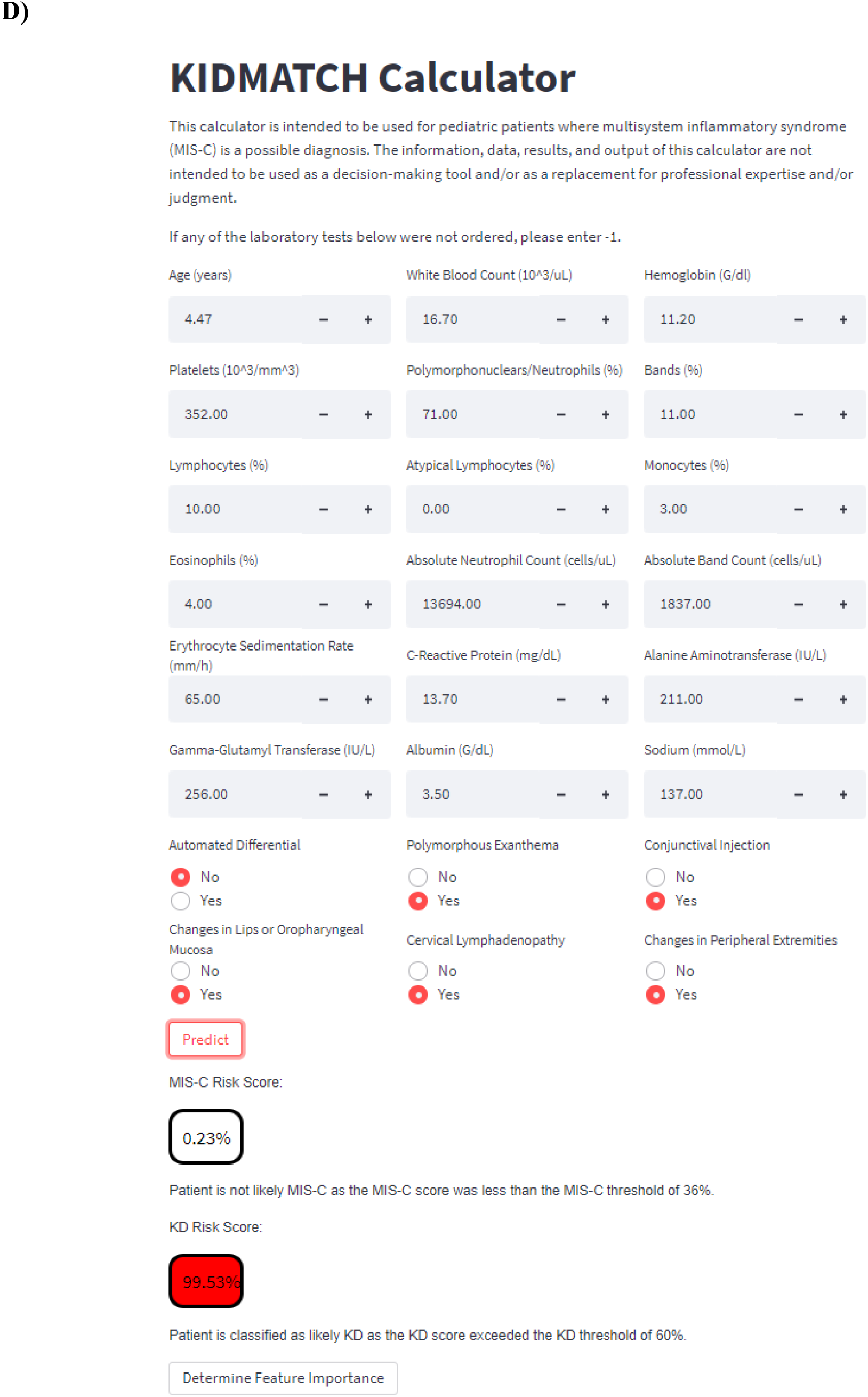
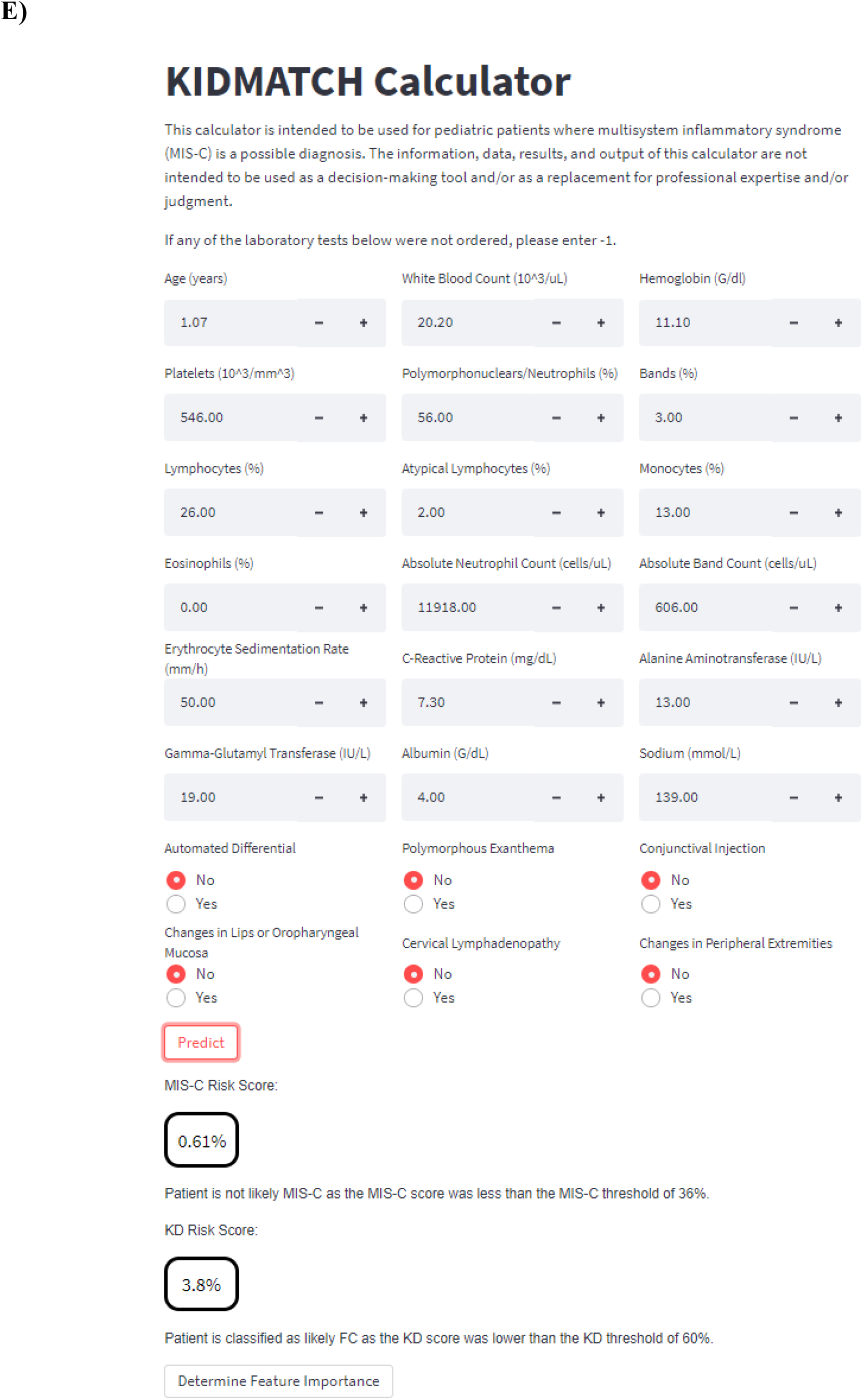
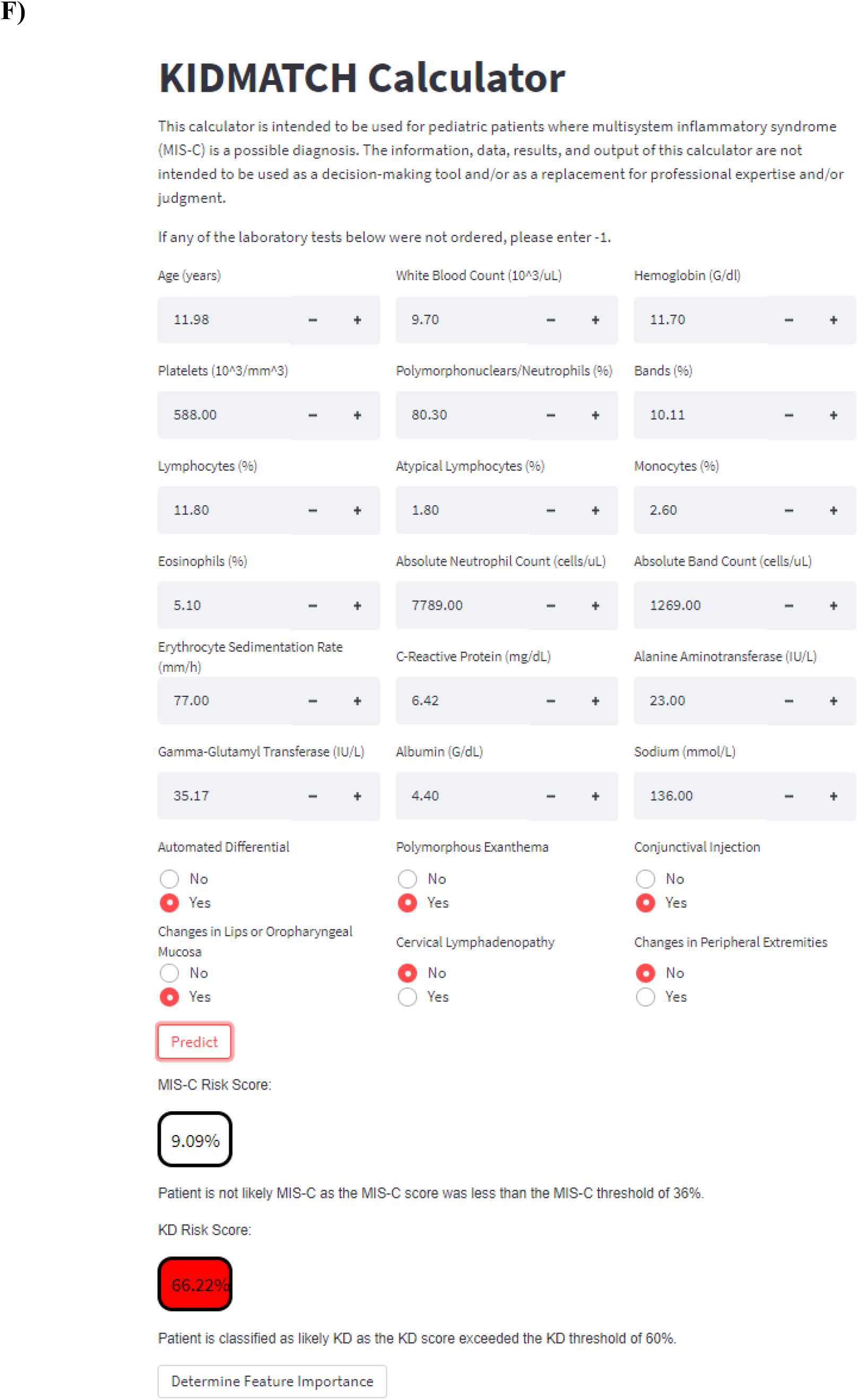
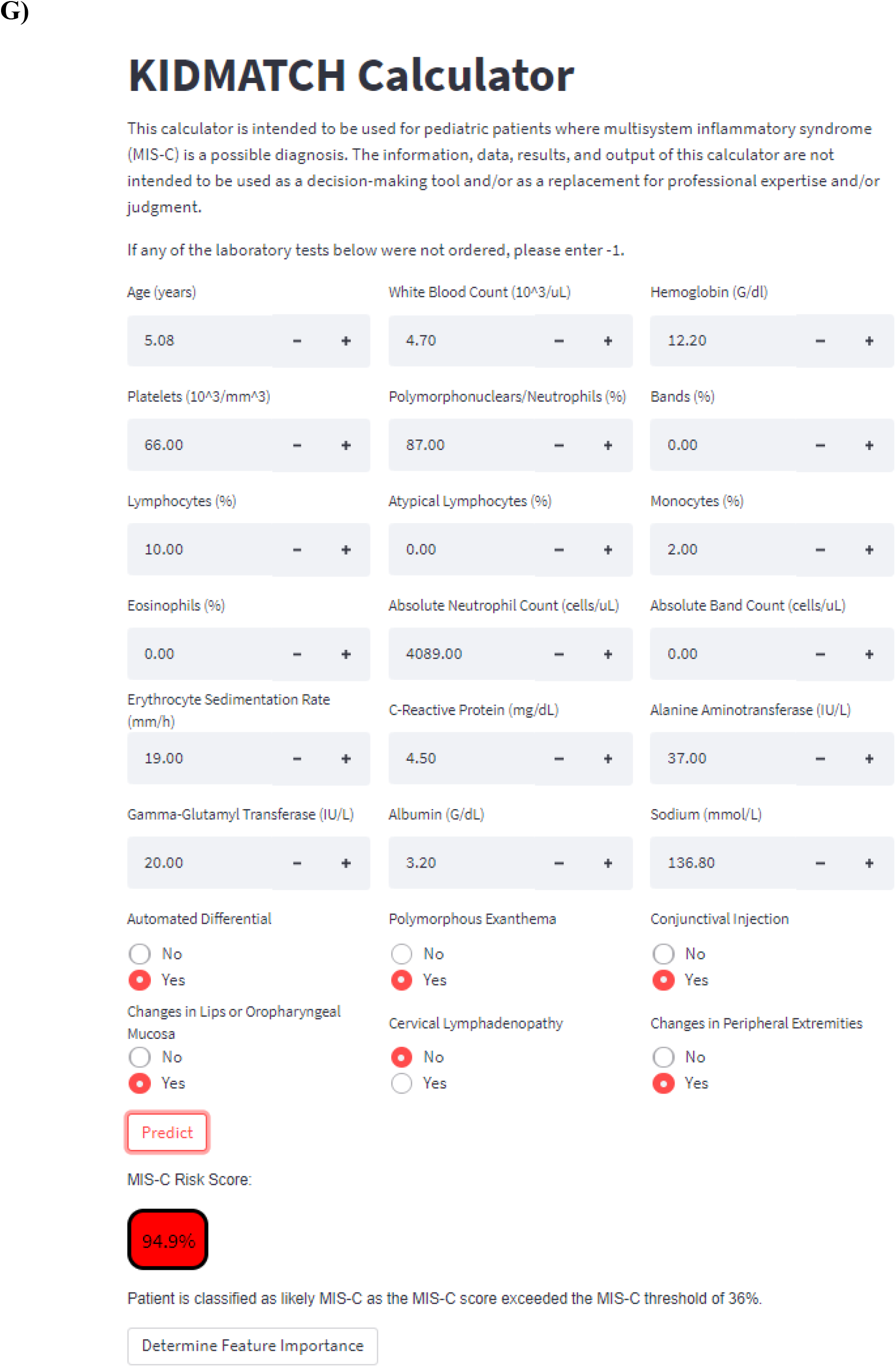
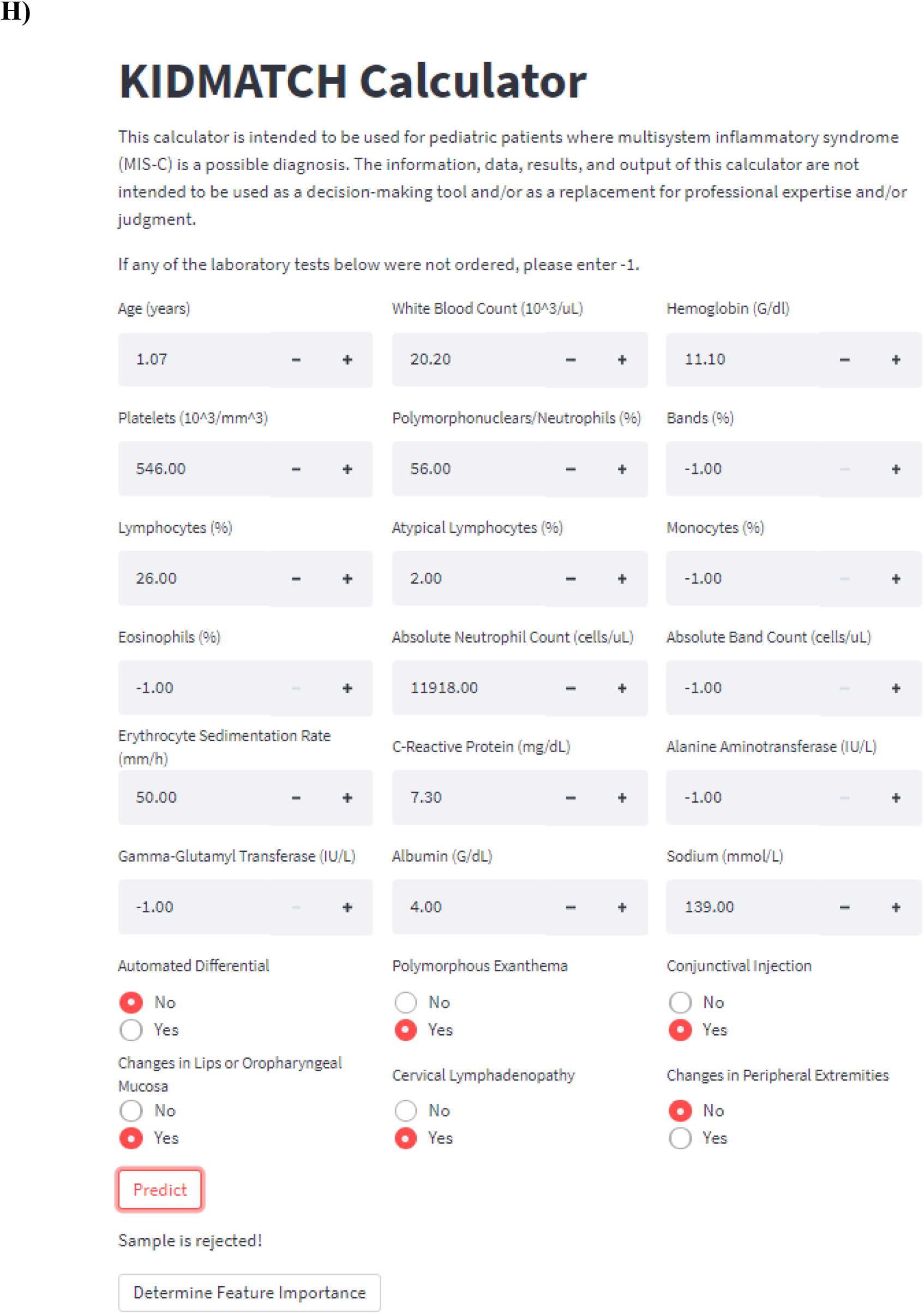
(A) User interface of the web-based calculator with values from a correctly classified MIS-C patient. Users enter the feature values, (B) predict risk scores, and (C) extract the important features contributing to the MIS-C risk score by pressing the “Predict” and “Feature Importance” buttons respectively. Other examples include risk scores for (D) a correctly classified KD patient, (E) a correctly classified FC patient, (F) a false negative patient (MIS-C classified as KD), (G) a false positive patient (KD classified as MIS-C), and (H) a patient rejected by the conformal prediction framework due to too many missing values.

**Supplementary Table 1:** Weighted F1 scores for missingness and risk score percentiles used to find the optimal combination for conformal prediction. The lowest percentile excluded (75^th^ percentile) was determined based on whether the included samples were greater than 200 for both the FC and KD cohort. 0.29% of samples had > 5 missing values.

| Percentile Excluded | Exclude data with > 1 missing value | Exclude data with > 2 missing values | Exclude data with > 3 missing values | Exclude data with > 4 missing values | Exclude data with > 5 missing values |
| --- | --- | --- | --- | --- | --- |
| 95 - 100 | **0.874** | 0.872 | 0.871 | 0.871 | 0.871 |
| 90 - 100 | 0.873 | 0.872 | 0.871 | 0.871 | 0.871 |
| 85 - 100 | 0.873 | 0.872 | 0.871 | 0.871 | 0.871 |
| 80 - 100 | 0.874 | 0.872 | 0.871 | 0.871 | 0.871 |
| 75 - 100 | 0.874 | 0.873 | 0.872 | 0.871 | 0.871 |

**Supplementary Table 2.** Number of subjects with missing data. Bolded features are features included in the model. LN: lymph nodes, LV: left ventricle, WBC: white blood cell count, Polys: polymorphonuclears or neutrophils, ANC: absolute neutrophil count, ABC: absolute band count, ALC: absolute lymphocyte count, ZHgb: hemoglobin concentration normalized for age, PLT: platelet count, ESR: erythrocyte sedimentation rate, CRP: C-reactive protein, ALT: Alanine aminotransferase, GGT: gamma-glutamyltransferase

|  | MIS-C<br>(n=131) | Kawasaki<br>disease<br>(n=775) | Febrile<br>control<br>(n=673) |
| --- | --- | --- | --- |
| **Age** | 0 | 0 | 0 |
| Male | 1 | 0 | 0 |
| Ethnicity | 0 | 0 | 0 |
| Median Zmax | 2 | 0 | 673 |
| Lowest LV Ejection Fraction | 2 | 5 | 673 |
| Illness day of Sample<br>collection | 0 | 0 | 0 |
| **Automated differential** | 0 | 0 | 0 |
| Clinical Sign |  |  |  |
| **Rash** | 0 | 0 | 0 |
| **Red Eyes** | 0 | 0 | 0 |
| **Oral Changes** | 0 | 0 | 0 |
| **Enlarged cervical LN** | 0 | 0 | 0 |
| **Extremity Changes** | 0 | 0 | 0 |
| Laboratory data |  |  |  |
| **WBC** | 0 | 0 | 0 |
| **Polys** | 0 | 0 | 1 |
| **Bands** | 29 | 92 | 111 |
| **Lymphocytes** | 0 | 0 | 1 |
| **Atypical lymphocytes** | 69 | 134 | 114 |
| **Monocytes** | 1 | 0 | 1 |
| **Eosinophil** | 0 | 10 | 7 |
| **ANC** | 0 | 0 | 1 |
| **ABC** | 29 | 92 | 111 |
| **ALC** | 0 | 0 | 1 |
| **ZHgb** | 0 | 0 | 0 |
| **PLT** | 0 | 1 | 2 |
| **ESR** | 10 | 12 | 31 |
| **CRP** | 1 | 6 | 19 |
| **ALT** | 1 | 5 | 43 |
| **GGT** | 66 | 14 | 71 |
| **Albumin** | 2 | 22 | 56 |

**Supplementary Table 3.**
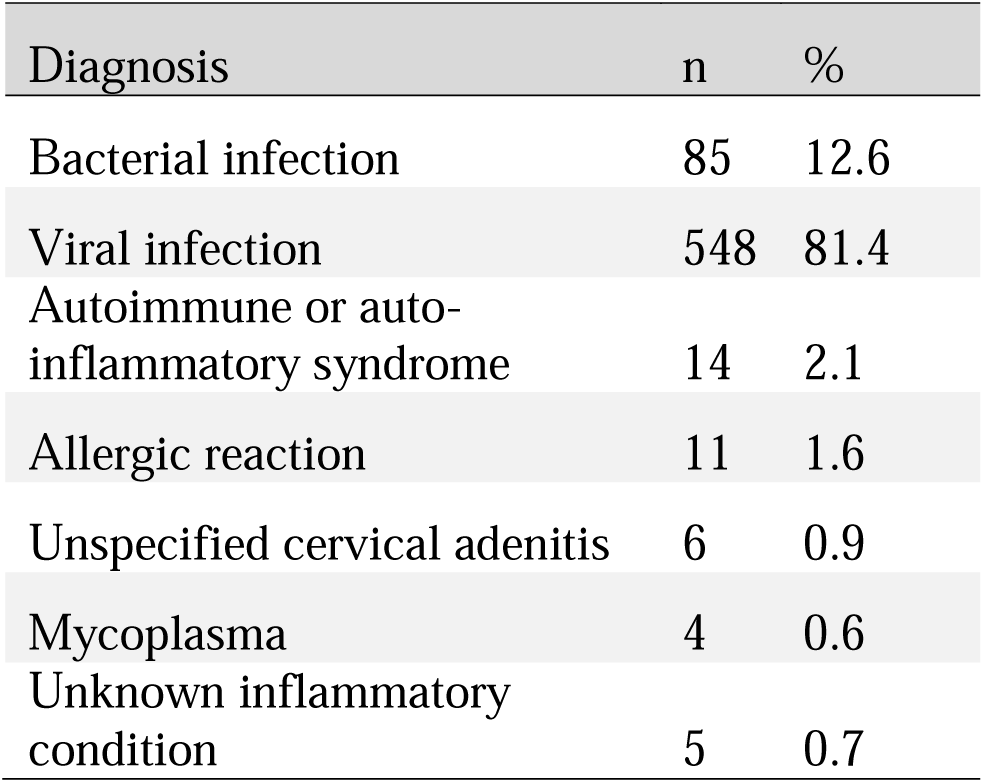
List of diagnoses for febrile children (FC). Subjects with bacterial infection included children with a positive culture from an otherwise sterile site, culture-proven bacterial enteritis, scarlet fever and toxic shock with appropriate supporting cultures and serology, and cervical lymphadenitis that responded to antibiotic therapy. Subjects with viral infection included children with PCR-proven specific viral diagnoses or children with “viral syndrome” defined as a self-limited illness that resolved without treatment and without apparent sequelae. All FC had fever for at least 3 days and at least one clinical criteria for KD.

**Supplementary Table 4:** Predicted classifications of MIS-C patients from the CHARMS Study Group by site number.

| MIS-C Subjects Predicted As |  |  |  |  |  |
| --- | --- | --- | --- | --- | --- |
| Site | FC | KD | MISC | Rejected | Total |
| 2 | 1 | 0 | 1 | 0 | 2 |
| 3 | 1 | 1 | 13 | 1 | 16 |
| 12 | 0 | 0 | 8 | 0 | 8 |
| 15 | 0 | 0 | 2 | 0 | 2 |
| 22 | 0 | 0 | 5 | 0 | 5 |
| 23 | 0 | 0 | 1 | 0 | 1 |
| 27 | 0 | 0 | 1 | 0 | 1 |
| 31 | 0 | 0 | 6 | 0 | 6 |
| 33 | 1 | 0 | 10 | 0 | 11 |
| 34 | 0 | 0 | 3 | 0 | 4 |
| 35 | 0 | 0 | 9 | 0 | 9 |
| 37 | 1 | 0 | 6 | 0 | 7 |
| 39 | 0 | 0 | 8 | 1 | 9 |
| 40 | 0 | 0 | 2 | 0 | 2 |

